# Phenotyping Neuropsychiatric Symptoms Profiles of Alzheimer’s Disease Using Cluster Analysis on EEG Power

**DOI:** 10.1101/2020.08.20.20179051

**Authors:** Friedrich Liu, Jong-Ling Fuh, Chung-Kang Peng, Albert C. Yang

**Affiliations:** Boston University Academy, Boston, MA, USA; Center for Dynamical Biomarkers, Beth Israel Deaconess Medical Center/Harvard Medical School, Boston, MA, USA; Division of General Neurology, Neurological Institute, Taipei Veterans General Hospital, Taipei, Taiwan; Faculty of Medicine, National Yang-Ming University, Taipei, Taiwan; Institute of Brain Sciences /Digital Medicine Center, National Yang-Ming University, Taipei, Taiwan

**Keywords:** Dementia, Electroencephalography (EEG), Behavioral and psychological symptoms, Cluster analysis, Fourier transform

## Abstract

There has been an increasing interest in studying electroencephalogram (EEG) as a biomarker of Alzheimer’s disease but the association between EEG signals and patients’ neuropsychiatric symptoms remains unclear. We studied EEG signals of patients with Alzheimer’s disease to explore the associations between patients’ neuropsychiatric symptoms and clusters of patients based on their EEG powers. A total of 69 patients with mild Alzheimer’s disease (the Clinical Dementia Rating = 1) were enrolled and their EEG signals from 19 channels/electrodes were recorded in three sessions for each patient. The EEG power was calculated by Fourier transform for the four frequency bands (beta: 13–40 Hz, alpha: 8–13 Hz, theta: 4–8 Hz, and delta: < 4 Hz). We performed K-means cluster analysis to classify the 69 patients into two distinct groups by the log-transformed EEG powers (4 frequency bands x 19 channels) for the three EEG sessions. In each session, both clusters were compared with each other to assess the differences in their behavioral/psychological symptoms in terms of the Neuropsychiatric Inventory (NPI) score. While EEG band powers were highly consistent across all three sessions before clustering, EEG band powers were different between the two clusters in each session, especially for the delta waves. The delta band powers differed significantly between the two clusters in most channels across the three sessions. Patients’ demographics and cognitive function were not different between both clusters. However, their behavioral/psychological symptoms were different between the two clusters classified based on EEG powers. A higher NPI score was associated with the clustering of higher EEG powers. The results suggest that EEG delta band power correlates to behavioral and psychological symptoms among patients with mild Alzheimer’s disease. The clustering approach of EEG signals may provide a novel and cost-effective method to differentiate the severity of neuropsychiatric symptoms and/or predict the prognosis for Alzheimer’s patients.

## 1. Introduction

Alzheimer’s disease is a type of neurodegenerative disorder which affects 5.8 million elderly people in the United States alone.[1] It is the sixth leading cause of death in the general population of the United States and is also the leading cause of disability and morbidity amongst the elderly.[1] As of 2019, the global percentage of people above the age of 60 who are affected by Alzheimer’s disease is estimated to be between 5% and 8%.[2] Symptoms of Alzheimer’s disease include loss of memory, reduced cognitive functions, and psychological conditions such as hallucination, behavioral and mood changes.[3,4]

Currently, dementia due to Alzheimer’s disease is diagnosed by a set of criteria involving patients’ cognitive function and neuropsychiatric history.[3,4] The assessment of Alzheimer’s disease includes the testing the patient’s memory, verbal skills, problem solving skills, thinking skills, and mood through a series of questionnaires. Close acquaintances of the patient are also questioned about the patient’s memory, cognitive ability, and recent behavior.[1] However, this method of diagnosis may not capture the heterogeneous nature of the disease. Recently, researchers have harbored significant interest in developing an objective method of classifying Alzheimer’s disease and assessing the severity of the condition that is based on biological markers.[5,6] Reliable quantitative assessments could offer great utility in clinical settings and could provide an alternative method for evaluating Alzheimer’s disease.[6–8] These assessments would be especially useful if they were simple and inexpensive to conduct.

Pathophysiologically, dementia of the Alzheimer type is caused by a build-up of beta-amyloid peptide in the brain, which in turn leads to synaptic dysfunction, impairing the vital cognitive functioning of the brain.[8–10] One of the convenient measurements that can assess brain functioning is the electroencephalography (EEG).[11–13] The EEG is a non-invasive, widely-available, and economical method that offers high temporal resolution, making it more attractive than functional magnetic resonance imaging (MRI) and positron-emission tomography (PET). Moreover, EEG scans directly measure neurological signaling while MRI and PET offer only hemodynamics and metabolic signals, respectively.[12]

While previous studies had examined the difference in EEG scans of Alzheimer’s patients versus non-Alzheimer’s patients, this study aimed to explore the potential difference in EEG scans of heterogeneous Alzheimer’s patients. Our hypothesis was that between patients with the same severity of disease, there would still be a difference in their neuropsychiatric symptoms with a corresponding difference in their EEG signals.

## 2. Material and methods

### 2.1 Participants

This study group was composed of 69 patients (42 women versus 27 men) with mild Alzheimer’s disease, 61 to 90 years of age (mean ± standard deviation, SD = 78.0 ± 6.7). All of the research participants were enrolled from the Dementia Clinic at the Neurological Institute of the Taipei Veterans General Hospital in Taiwan. The diagnosis for Alzheimer’s disease was based on the criteria of the National Institute of Neurological and Communicative Disorders and the Stroke/Alzheimer’s Disease and Related Disorders Association.[4] In light of the study purpose to assess EEG signals among patients with similar severity of dementia, only patients characterized as having mild Alzheimer’s dementia (i.e. Clinical Dementia Rating, CDR scale = 1)[14] were included in this study.

The cognitive functioning of the patients was evaluated using the Mini-Mental State Examination (MMSE),[15] a verbal category fluency test, and the forward-and-backward digit span tasks subset of the Wechsler Adult Intelligence Scale. Their behavioral and psychological symptoms were evaluated using the Neuropsychiatric Inventory (NPI), a quantitative assessment of over 12 neuropsychiatric domains: delusions, hallucinations, dysphoria, anxiety, agitation/aggression, euphoria, disinhibition, irritability/lability, apathy, aberrant motor activity, appetite, and night-time behavior disturbances.[16]

### 2.2 EEG signal processing

All patients received digital EEG recording in the EEG examination room and the details of the EEG recording protocol has been previously reported.[17] Each patient had three separate sessions of resting eye-closed EEG scan for 10 to 20 seconds. EEG signals from the 19 electrodes (Fp1, Fp2, F7, F3, Fz, F4, F8, T3, C3,Cz, C4, T4, T5, P3, Pz, P4, T6, O1, and O2) were recorded according to the standard 10–20 system with reference to linked earlobe electrodes at a sampling rate of 256 Hz. Initial filter settings were low-pass filter frequency of 70 Hz, high-pass filter of 0.05 Hz, notch filter of 60 Hz, and electrode impedances below 3 kΩ. A technician monitored the process and would alert the patient if signs of drowsiness/sleep appeared.[17]

For the EEG scan from each electrode per session with eyes closed, a 10-second segment of artifact-free EEG signals was manually extracted after careful visual inspection.[17] Each patient had three EEG segments of 10 seconds each from each electrode, and these EEG data were used for the subsequent analyses. The EEG signal processing and analysis in this study were all done on the Matrix Laboratory (MATLAB) software (MathWorks, Inc., Natick, Massachusetts, United States).

The original EEG data from each electrode per segment as a function of voltage over time were transformed using the Fourier transform to yield the Welch’s periodogram of the power spectral density versus frequency. The EEG signals were classified by the four conventional frequency bands defined as beta (13–40 Hz), alpha (8–13 Hz), theta (4–8 Hz), and delta (< 4 Hz). The absolute EEG power was calculated by integrating the power spectral density with respect to frequency for the corresponding frequency band. Each patient has 76 band powers since each of the 19 electrodes has 4 frequency bands. The EEG band powers were then log-transformed and a 69 (patients) by 76 (log-transformed band powers) result matrix was constructed for each of the three segments.

### 2.3 Cluster analysis

Cluster analysis was performed on each of the three result matrices of log-transformed band powers using k-means clustering on MATLAB to classify the 69 patients in each of the three segments into two distinct clusters, making 6 overall clusters. The similarities between clusters were evaluated in terms of the Jaccard similarity coefficient, a measure of intersection over union of the groups compared.

### 2.4 Statistical analysis

For each of the three EEG segments, both clusters generated by K-means method on their EEG powers were compared with each other to assess the differences in their demographics, cognitive functioning, and neuropsychiatric symptoms. The demographics of the patients included age, sex, education, and duration of illness. The cognitive functioning included MMSE, verbal fluency, digit forward span test, and digit backward span test. The NPI variables contained the 12 neuropsychiatric domains listed above with higher scores indicating worse conditions. Independent-sample t-tests were conducted to compare the continuous variables between both clusters whereas the chi-square tests were used to compare the categorical variables such as gender distribution. Two-sided p values of less than 0.05 were considered statistically significant. IBM SPSS Statistics, version 25.0 (SPSS Inc., Chicago, Illinois, United States) was used for all of the statistical analyses.

## 3. Results

### 3.1 Scalp EEG power features of patients with dementia

The absolute EEG power spectral density analysis was performed on the EEG data among the three segments of the 69 patients with mild Alzheimer’s disease. Topographic plots showed the EEG power features mapping the EEG band power for the 19 electrodes in a 2-dimensional circular view looking down from the top of the head (Figure 1). Figure 1A shows that the EEG power features for the four frequency bands were highly consistent across all three segments before clustering. For the delta frequency band, the EEG signals from left frontal electrodes (FP1) were strongest across all three segments. In contrast, for the theta or alpha frequency band, the EEG powers were weak globally across all three segments.

**Figure 1:**
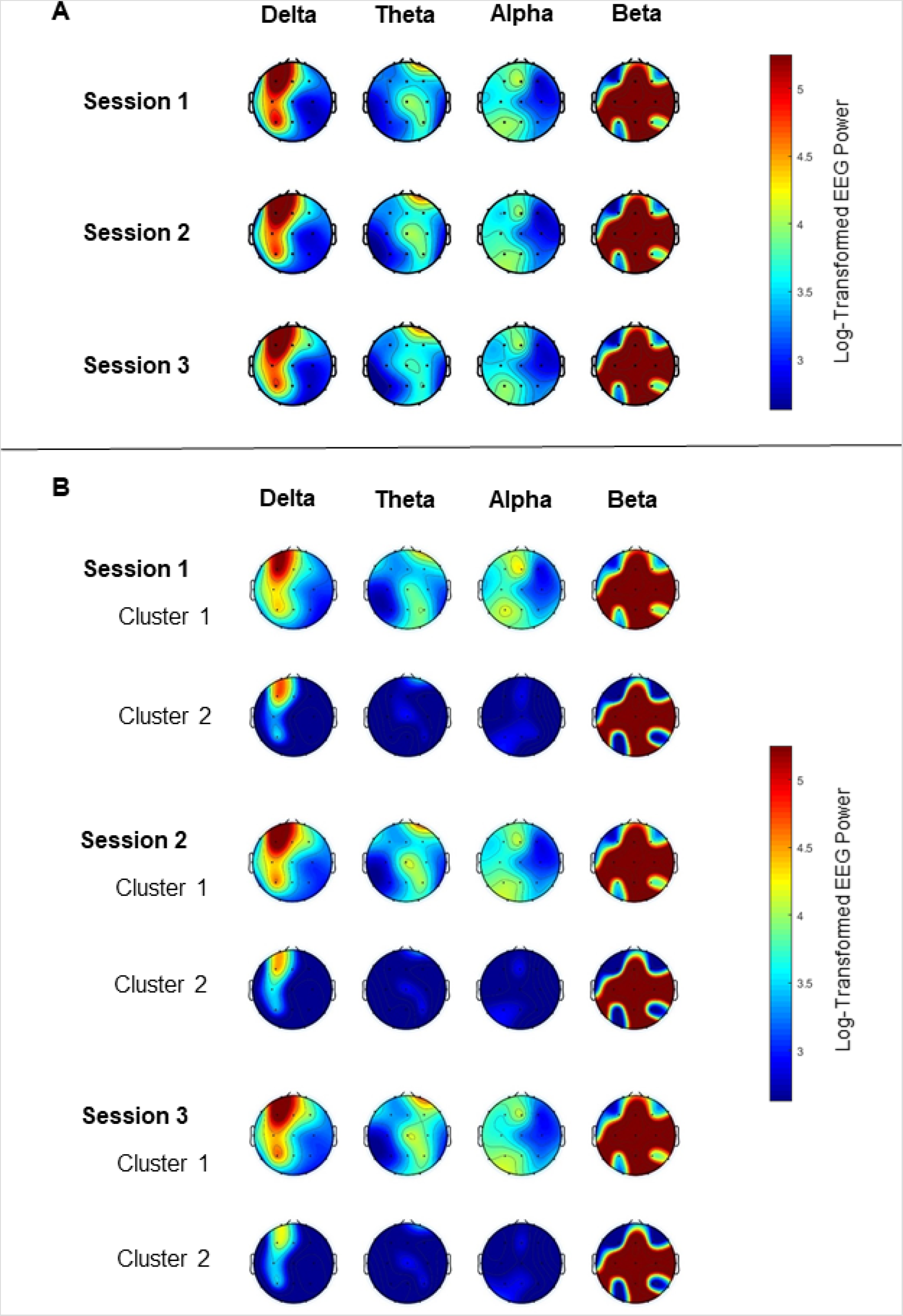
Topography of EEG power for the four frequency bands among the three EEG sessions. The color spectrum corresponds to the log-transformed EEG power for the delta (< 4 Hz), theta (4–8 Hz), alpha (8–13 Hz) and beta (13–40 Hz) band. Panel A shows that EEG band powers are consistent across all three sessions. Panel B shows that EEG band powers are different between the two clusters in each of the three sessions, especially for the delta frequency band.

### 3.2 Cluster analysis on EEG powers

For each of the three EEG segments, cluster analysis was performed to classify the 69 patients into two distinct clusters based on their EEG frequency band powers. Clusters 1 and 2 contained 33 vs. 36, 33 vs. 36, and 34 vs. 35 patients for segment 1, segment 2, and segment 3, respectively. Table 1 shows the similarities in clustering between different segments in terms of the Jaccard similarity index which measures the intersection over union of the clusters compared. Overall, the clustering patterns between the three segments were highly consistent as the Jaccard index ranged from 73.7% to 77.5%, significantly above the value of around 31% by random grouping. Figure 1B also reveals the consistency of clustering on EEG band powers across the three segments.

**Table 1.** Comparisons of the clustering in terms of the Jaccard index (%) for the cluster analysis on EEG powers between the three EEG sessions.

|  |  | Session 1 |  | Session 2 |  | Session 3 |  |
| --- | --- | --- | --- | --- | --- | --- | --- |
|  |  | Cluster 1 | Cluster 2 | Cluster 1 | Cluster 2 | Cluster 1 | Cluster 2 |
| Session 1 | Cluster 1 | - | 0 |  |  |  |  |
|  | Cluster 2 | 0 | - |  |  |  |  |
| Session 2 | Cluster 1 | 73.7 | 7.8 | - | 0 |  |  |
|  | Cluster 2 | 7.8 | 75.6 | 0 | - |  |  |
| Session 3 | Cluster 1 | 76.3 | 7.7 | 76.3 | 7.7 | - | 0 |
|  | Cluster 2 | 6.3 | 77.5 | 6.3 | 77.5 | 0 | - |

In each of the three segments, the two clusters generated by K-means clustering methods were distinctive with each other regarding their EEG power, particularly for the delta frequency band (Figure 1B). For every segment, the delta band power, theta band power, and alpha band power of one cluster (cluster 1) appeared stronger than that for the corresponding electrode of the other cluster (cluster 2). Table 2 shows that the delta band powers differed significantly between the two clusters in most of the 19 scalp electrodes across the three sessions. Although the strongest signals came from the left frontal area (Fp1), the EEG delta band power for Fp1 of cluster 1 was still greater than that of cluster 2.

**Table 2.**
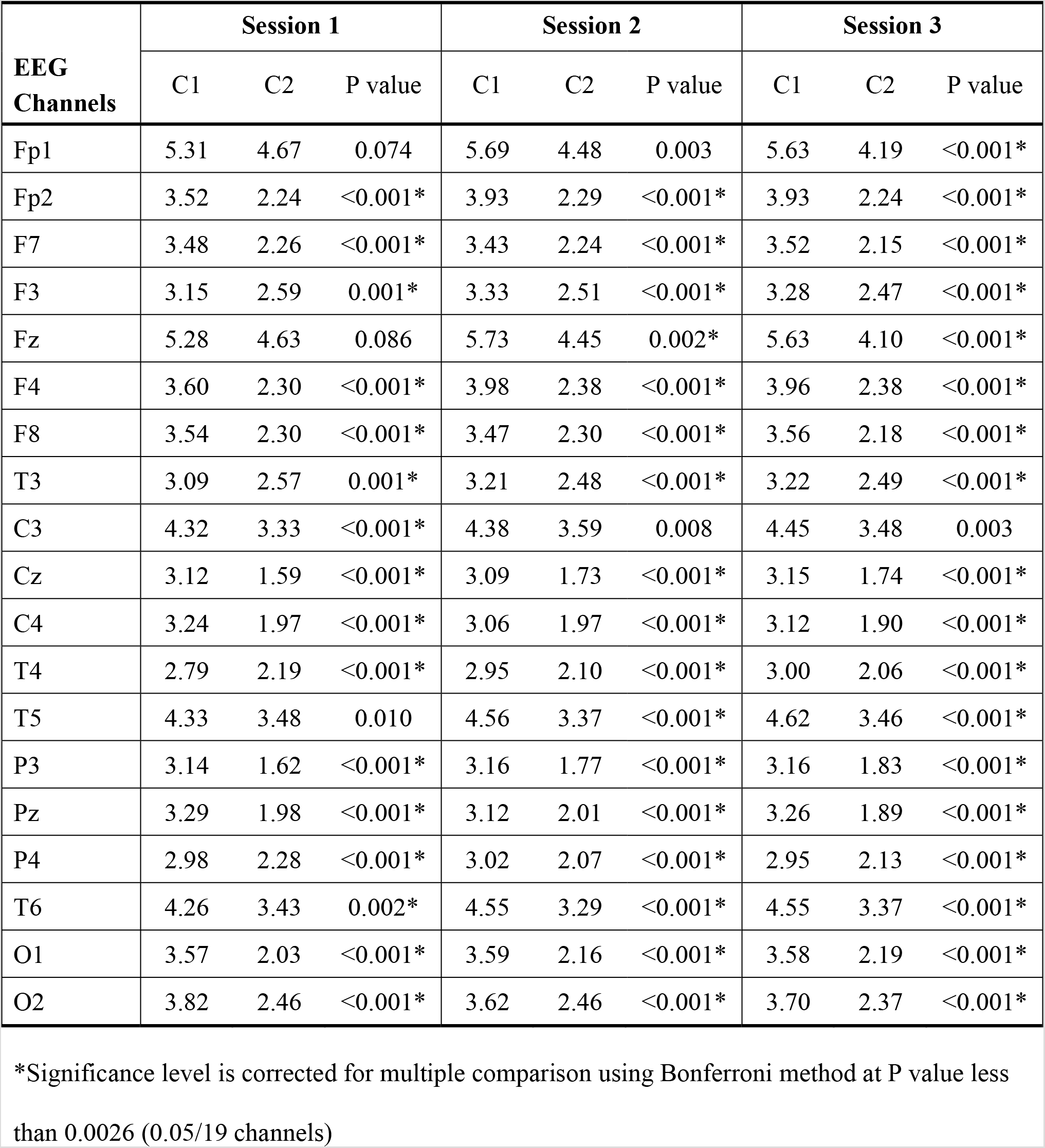
Distinction in log-transformed EEG delta band power between both clusters across 19 EEG channels among the three sessions.

| EEG Channels | Session 1 |  |  | Session 2 |  |  | Session 3 |  |  |
| --- | --- | --- | --- | --- | --- | --- | --- | --- | --- |
|  | C1 | C2 | P value | C1 | C2 | P value | C1 | C2 | P value |
| Fp1 | 5.31 | 4.67 | 0.074 | 5.69 | 4.48 | 0.003 | 5.63 | 4.19 | <0.001* |
| Fp2 | 3.52 | 2.24 | <0.001* | 3.93 | 2.29 | <0.001* | 3.93 | 2.24 | <0.001* |
| F7 | 3.48 | 2.26 | <0.001* | 3.43 | 2.24 | <0.001* | 3.52 | 2.15 | <0.001* |
| F3 | 3.15 | 2.59 | 0.001* | 3.33 | 2.51 | <0.001* | 3.28 | 2.47 | <0.001* |
| Fz | 5.28 | 4.63 | 0.086 | 5.73 | 4.45 | 0.002* | 5.63 | 4.10 | <0.001* |
| F4 | 3.60 | 2.30 | <0.001* | 3.98 | 2.38 | <0.001* | 3.96 | 2.38 | <0.001* |
| F8 | 3.54 | 2.30 | <0.001* | 3.47 | 2.30 | <0.001* | 3.56 | 2.18 | <0.001* |
| T3 | 3.09 | 2.57 | 0.001* | 3.21 | 2.48 | <0.001* | 3.22 | 2.49 | <0.001* |
| C3 | 4.32 | 3.33 | <0.001* | 4.38 | 3.59 | 0.008 | 4.45 | 3.48 | 0.003 |
| Cz | 3.12 | 1.59 | <0.001* | 3.09 | 1.73 | <0.001* | 3.15 | 1.74 | <0.001* |
| C4 | 3.24 | 1.97 | <0.001* | 3.06 | 1.97 | <0.001* | 3.12 | 1.90 | <0.001* |
| T4 | 2.79 | 2.19 | <0.001* | 2.95 | 2.10 | <0.001* | 3.00 | 2.06 | <0.001* |
| T5 | 4.33 | 3.48 | 0.010 | 4.56 | 3.37 | <0.001* | 4.62 | 3.46 | <0.001* |
| P3 | 3.14 | 1.62 | <0.001* | 3.16 | 1.77 | <0.001* | 3.16 | 1.83 | <0.001* |
| Pz | 3.29 | 1.98 | <0.001* | 3.12 | 2.01 | <0.001* | 3.26 | 1.89 | <0.001* |
| P4 | 2.98 | 2.28 | <0.001* | 3.02 | 2.07 | <0.001* | 2.95 | 2.13 | <0.001* |
| T6 | 4.26 | 3.43 | 0.002* | 4.55 | 3.29 | <0.001* | 4.55 | 3.37 | <0.001* |
| O1 | 3.57 | 2.03 | <0.001* | 3.59 | 2.16 | <0.001* | 3.58 | 2.19 | <0.001* |
| O2 | 3.82 | 2.46 | <0.001* | 3.62 | 2.46 | <0.001* | 3.70 | 2.37 | <0.001* |

\*Significance level is corrected for multiple comparison using Bonferroni method at P value less than 0.0026 (0.05/19 channels)

### 3.3 Comparisons of demographic and clinical variables between clusters on EEG powers

Following cluster analysis on EEG powers, the two clusters generated by K-means method were compared with each other regarding demographic and clinical variables. In general, for each of the EEG segments, patients’ demographics and cognitive functions were not different between both clusters except for segment 1, which had more females in cluster 1 than in cluster 2 (75.8% vs. 47.2%; P = 0.015).

Regarding patients’ neuropsychiatric symptoms, Table 3 shows a clear trend that in all three segments, patients of cluster 1 exhibited higher NPI scores, total score or subdomain score, than cluster 2, indicating that the clustering of higher EEG powers may be associated with severer neuropsychiatric symptoms. For segment 1, the total NPI score for the first cluster was significantly higher than the second cluster (19.3 ± 18.6 vs. 10.0 ± 9.7; P = 0.014). For segment 2, the total NPI score for the first cluster was 17.2 ± 18.9 while the second cluster had a score of 11.9 ± 10.6. And for segment 3, the total NPI score for cluster 1 was 17.7 ± 18.7 while cluster 2 was 11.3 ± 10.3 (P = 0.086, Table 3).

**Table 3.**
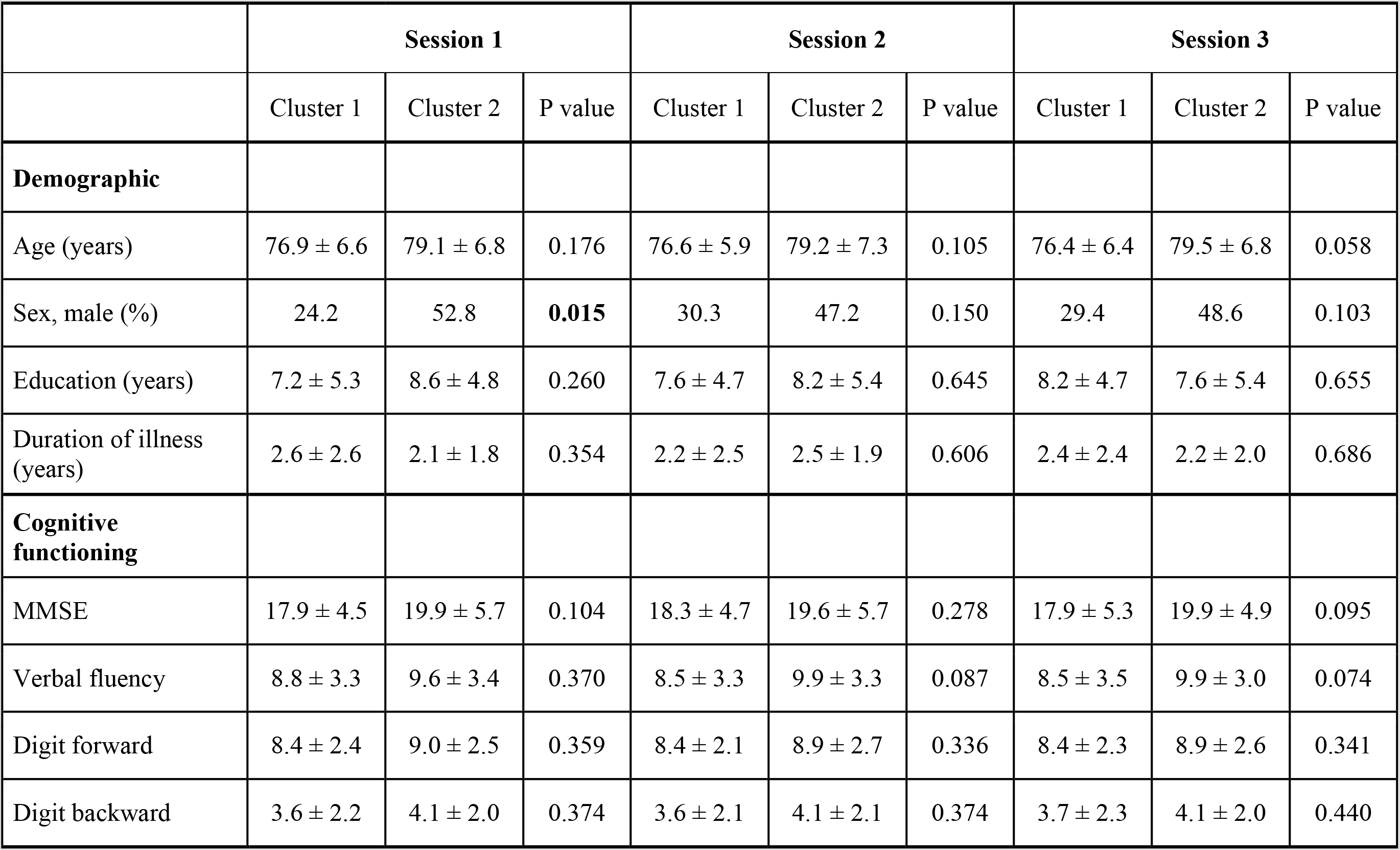
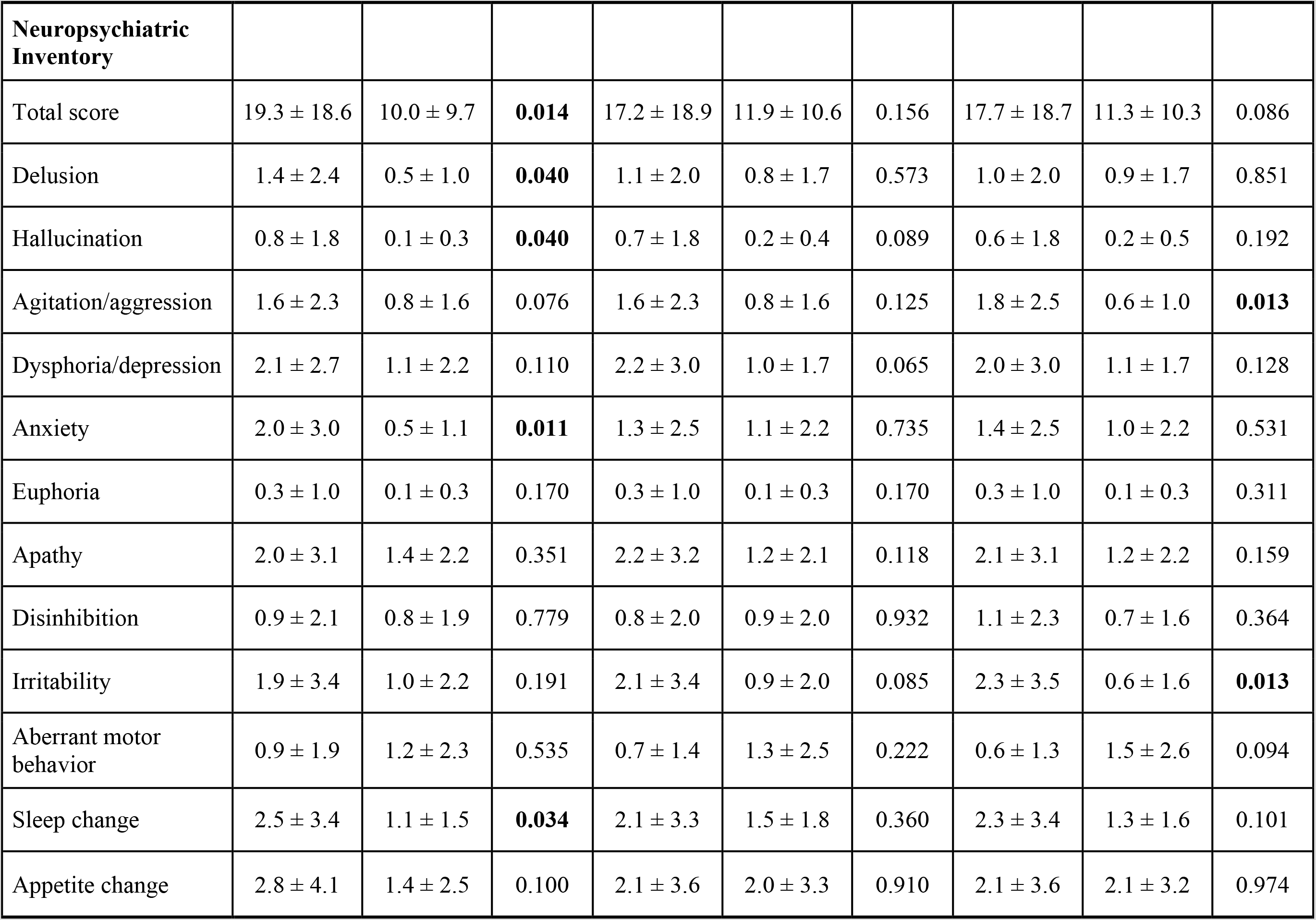
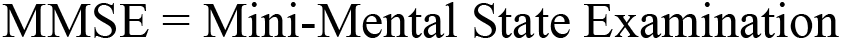
Comparison of cognition function and behavioral/psychiatric symptoms between both clusters of patients classified by their EEG powers for the three EEG sessions.

|  | Session 1 |  |  | Session 2 |  |  | Session 3 |  |  |
| --- | --- | --- | --- | --- | --- | --- | --- | --- | --- |
|  | Cluster 1 | Cluster 2 | P value | Cluster 1 | Cluster 2 | P value | Cluster 1 | Cluster 2 | P value |
| **Demographic** |  |  |  |  |  |  |  |  |  |
| Age (years) | 76.9 ± 6.6 | 79.1 ± 6.8 | 0.176 | 76.6 ± 5.9 | 79.2 ± 7.3 | 0.105 | 76.4 ± 6.4 | 79.5 ± 6.8 | 0.058 |
| Sex, male (%) | 24.2 | 52.8 | **0.015** | 30.3 | 47.2 | 0.150 | 29.4 | 48.6 | 0.103 |
| Education (years) | 7.2 ± 5.3 | 8.6 ± 4.8 | 0.260 | 7.6 ± 4.7 | 8.2 ± 5.4 | 0.645 | 8.2 ± 4.7 | 7.6 ± 5.4 | 0.655 |
| Duration of illness (years) | 2.6 ± 2.6 | 2.1 ± 1.8 | 0.354 | 2.2 ± 2.5 | 2.5 ± 1.9 | 0.606 | 2.4 ± 2.4 | 2.2 ± 2.0 | 0.686 |
| **Cognitive functioning** |  |  |  |  |  |  |  |  |  |
| MMSE | 17.9 ± 4.5 | 19.9 ± 5.7 | 0.104 | 18.3 ± 4.7 | 19.6 ± 5.7 | 0.278 | 17.9 ± 5.3 | 19.9 ± 4.9 | 0.095 |
| Verbal fluency | 8.8 ± 3.3 | 9.6 ± 3.4 | 0.370 | 8.5 ± 3.3 | 9.9 ± 3.3 | 0.087 | 8.5 ± 3.5 | 9.9 ± 3.0 | 0.074 |
| Digit forward | 8.4 ± 2.4 | 9.0 ± 2.5 | 0.359 | 8.4 ± 2.1 | 8.9 ± 2.7 | 0.336 | 8.4 ± 2.3 | 8.9 ± 2.6 | 0.341 |
| Digit backward | 3.6 ± 2.2 | 4.1 ± 2.0 | 0.374 | 3.6 ± 2.1 | 4.1 ± 2.1 | 0.374 | 3.7 ± 2.3 | 4.1 ± 2.0 | 0.440 |

| **Neuropsychiatric Inventory** |  |  |  |  |  |  |  |  |  |
| --- | --- | --- | --- | --- | --- | --- | --- | --- | --- |
| Total score | 19.3 ± 18.6 | 10.0 ± 9.7 | **0.014** | 17.2 ± 18.9 | 11.9 ± 10.6 | 0.156 | 17.7 ± 18.7 | 11.3 ± 10.3 | 0.086 |
| Delusion | 1.4 ± 2.4 | 0.5 ± 1.0 | **0.040** | 1.1 ± 2.0 | 0.8 ± 1.7 | 0.573 | 1.0 ± 2.0 | 0.9 ± 1.7 | 0.851 |
| Hallucination | 0.8 ± 1.8 | 0.1 ± 0.3 | **0.040** | 0.7 ± 1.8 | 0.2 ± 0.4 | 0.089 | 0.6 ± 1.8 | 0.2 ± 0.5 | 0.192 |
| Agitation/aggression | 1.6 ± 2.3 | 0.8 ± 1.6 | 0.076 | 1.6 ± 2.3 | 0.8 ± 1.6 | 0.125 | 1.8 ± 2.5 | 0.6 ± 1.0 | **0.013** |
| Dysphoria/depression | 2.1 ± 2.7 | 1.1 ± 2.2 | 0.110 | 2.2 ± 3.0 | 1.0 ± 1.7 | 0.065 | 2.0 ± 3.0 | 1.1 ± 1.7 | 0.128 |
| Anxiety | 2.0 ± 3.0 | 0.5 ± 1.1 | **0.011** | 1.3 ± 2.5 | 1.1 ± 2.2 | 0.735 | 1.4 ± 2.5 | 1.0 ± 2.2 | 0.531 |
| Euphoria | 0.3 ± 1.0 | 0.1 ± 0.3 | 0.170 | 0.3 ± 1.0 | 0.1 ± 0.3 | 0.170 | 0.3 ± 1.0 | 0.1 ± 0.3 | 0.311 |
| Apathy | 2.0 ± 3.1 | 1.4 ± 2.2 | 0.351 | 2.2 ± 3.2 | 1.2 ± 2.1 | 0.118 | 2.1 ± 3.1 | 1.2 ± 2.2 | 0.159 |
| Disinhibition | 0.9 ± 2.1 | 0.8 ± 1.9 | 0.779 | 0.8 ± 2.0 | 0.9 ± 2.0 | 0.932 | 1.1 ± 2.3 | 0.7 ± 1.6 | 0.364 |
| Irritability | 1.9 ± 3.4 | 1.0 ± 2.2 | 0.191 | 2.1 ± 3.4 | 0.9 ± 2.0 | 0.085 | 2.3 ± 3.5 | 0.6 ± 1.6 | **0.013** |
| Aberrant motor behavior | 0.9 ± 1.9 | 1.2 ± 2.3 | 0.535 | 0.7 ± 1.4 | 1.3 ± 2.5 | 0.222 | 0.6 ± 1.3 | 1.5 ± 2.6 | 0.094 |
| Sleep change | 2.5 ± 3.4 | 1.1 ± 1.5 | **0.034** | 2.1 ± 3.3 | 1.5 ± 1.8 | 0.360 | 2.3 ± 3.4 | 1.3 ± 1.6 | 0.101 |
| Appetite change | 2.8 ± 4.1 | 1.4 ± 2.5 | 0.100 | 2.1 ± 3.6 | 2.0 ± 3.3 | 0.910 | 2.1 ± 3.6 | 2.1 ± 3.2 | 0.974 |

MMSE = Mini-Mental State Examination

Specifically, for segment 1, patients in cluster 1 experienced a significantly higher level of delusion, hallucination, anxiety, and sleep change than patients in cluster 2. For segment 3, patients in cluster 1 experienced a significantly higher level of agitation/aggression and irritability than patients in cluster 2.

## 4. Discussion

In light of the goal to develop an objective method of classifying Alzheimer’s disease and assessing the severity of the neuropsychiatric conditions using EEG markers, we performed cluster analysis to categorize the Alzheimer’s patients into two groups based on their EEG band powers from 19 scalp electrodes. We found a clear linkage between the clustering on EEG powers and differences in the behavioral and psychological symptoms that a higher NPI score was associated with the clustering of higher EEG band powers, supporting the hypothesis that the discrepancy in neuropsychiatric symptoms of patients with the same severity of Alzheimer’s dementia would correspond to a difference in their EEG signals.

Although previous studies repeatedly demonstrated that patients with Alzheimer’s disease show an increase in delta/theta band power, a decline in alpha/beta band power, reduced complexity and impaired synchrony in EEG signals compared to non-Alzheimer individuals,[11,12] there have been limited data on assessing the EEG signals associated with neuropsychiatric conditions for Alzheimer’s patients with the same severity of dementia. While neuropsychiatric symptoms generally increase as the stage of dementia progresses, many symptoms still develop in the early stage of Alzheimer’s disease.[18–20] Moreover, our findings verified that Alzheimer’s patients may be heterogeneous regarding their behavioral and psychological symptoms even though they have the same severity of dementia.

Our finding of the association of neuropsychiatric symptoms with the clustering on patients’ EEG band powers suggests that the clustering approach on EEG signals may be a reliable and effective biomarker to characterize neuropsychiatric conditions and/or prognosis for Alzheimer’s patients with similar cognitive functions. Specifically, such a cost-effective approach may identify patients with increased odds of delusion, hallucination, anxiety, sleep change, irritability, and agitation/aggression, which should in turn inform the prognosis, organize preventive measures and appropriate care, and facilitate applicable treatment plans for these patients.[20]

Taking the clustering approach on absolute EEG band powers provide several advantages. First, it is safe, convenient, and cost-effective. Second, the results of the clustering seemed stable and consistent that were displayed in our study. Moreover, as expected, the clustering was not associated with the demographics or cognitive functioning of Alzheimer’s patients with the same severity of disease. Third, without predetermined constraints, EEG signals for the four frequency bands from all 19 scalp electrodes, instead of information from only single band or local channels, were considered for the cluster analysis. Nevertheless, we found that the alpha band power, theta band power, and especially delta band power were grossly different between the two clusters in almost all channels and the clustering of higher EEG band powers corresponded to the group of patients with more severe neuropsychiatric conditions. Although we did not find prior research using the same approach that could corroborate our findings, previous studies reported that the changes of EEG signals develop gradually and the increase in EEG delta band power typically occurs in later stages across the Alzheimer’s disease continuum.[12,21,22]

Even though our findings shed light on the development of a novel model for phenotyping neuropsychiatric symptoms profiles of Alzheimer’s disease, there are still a few limitations to be noted. First, the major limitation is the relatively small number of participants which leads to insufficient statistical power to demonstrate the statistically significant differences in NPI scores between the two clusters for every EEG session. However, the trends showing that a higher NPI is associated with the clustering of higher EEG powers were apparent. Second, this is a cross-sectional study. Future large-scale prospective studies are warranted to investigate the long-term clinical outcomes of Alzheimer’s patients and to corroborate our findings.

In summary, the neuropsychiatric symptoms were different between the two groups of patients with the same severity of Alzheimer’s dementia categorized by cluster analysis based on EEG powers. A higher NPI score was associated with the clustering of higher EEG powers, particularly in delta frequency band. The results of the study suggested that EEG band powers correlate to behavioral/psychological symptoms among patients with Alzheimer’s disease. The clustering approach of EEG signals may provide a novel and cost-effective method to differentiate the severity of neuropsychiatric symptoms and/or predict the prognosis for Alzheimer’s patients.

## Data Availability

The electroencephalogram (EEG) data involved in the manuscript is an secondary analysis from the published data in the paper "Cognitive and neuropsychiatric correlates of EEG dynamic complexity in patients with Alzheimer's Disease" Progress in Neuro-Psychopharmacology & Biological Psychiatry 47 (2013) 52-61.

## Acknowledgments

Friedrich Liu was supported by the Center for Dynamical Biomarkers at Beth Israel Deaconess Medical Center/Harvard Medical School, Boston, MA, USA. Dr. Albert C. Yang was supported by grants [109–2628-B-010–011; 109–2321-B-010–006] from the Ministry of Science and Technology, Taiwan. Dr. Albert C. Yang was also supported by Mt. Jade Young Scholarship Award from the Ministry of Education of Taiwan as well as the Brain Research Center, National Yang-Ming University and the Ministry of Education (Aim for the Top University Plan), Taipei, Taiwan. Dr. Jong-Ling Fuh was supported by the Ministry of Science and Technology of Taiwan (109–2314-B-075–052-MY2) and Taipei Veterans General Hospital (V109C-061 and VGHUST109-V1–5–1).

## Notes

### Competing Interest Statement

The authors have declared no competing interest.

### Author Declarations

Our study was approved by the Institutional Review Board of Taipei Veterans General Hospital to conduct retrospective analysis of the patients' clinical and EEG data.

